# Myofascial Frequency Syndrome: A novel syndrome of bothersome lower urinary tract symptoms associated with myofascial pelvic floor dysfunction

**DOI:** 10.1101/2023.04.14.23288590

**Authors:** A. Lenore Ackerman, Nicholas J. Jackson, Ashley T. Caron, Melissa R. Kaufman, Jonathan C. Routh, Jerry L. Lowder

## Abstract

**Background:** Patients presenting with lower urinary tract symptoms (LUTS) are historically classified to several symptom clusters, primarily overactive bladder (OAB) and interstitial cystitis/bladder pain syndrome (IC/BPS). Accurate diagnosis, however, is challenging due to overlapping symptomatic features, and many patients do not readily fit into these categories. To enhance diagnostic accuracy, we previously described an algorithm differentiating OAB from IC/BPS. Herein, we sought to validate the utility of this algorithm for identifying and classifying a real-world population of individuals presenting with OAB and IC/BPS and characterize patient subgroups outside the traditional LUTS diagnostic paradigm.

**Methods:** An *Exploratory cohort* of 551 consecutive female subjects with LUTS evaluated in 2017 were administered 5 validated genitourinary symptom questionnaires. Application of the LUTS diagnostic algorithm classified subjects into controls, IC/BPS, and OAB, with identification of a novel group of highly bothered subjects lacking pain or incontinence. Symptomatic features of this group were characterized by statistically significant differences from the OAB, IC/BPS and control groups on questionnaires, comprehensive review of discriminate pelvic exam, and thematic analysis of patient histories. In a *Reassessment cohort* of 215 subjects with known etiologies of their symptoms (OAB, IC/BPS, asymptomatic microscopic hematuria, or myofascial dysfunction confirmed with electromyography), significant associations with myofascial dysfunction were identified in a multivariable regression model. Pre-referral and specialist diagnoses for subjects with myofascial dysfunction were catalogued.

**Findings:** Application of a diagnostic algorithm to an unselected group of 551subjects presenting for urologic care identified OAB and IC/BPS in 137 and 96 subjects, respectively. An additional 110 patients (20%) with bothersome urinary symptoms lacked either bladder pain or urgency characteristic of IC/BPS and OAB, respectively. In addition to urinary frequency, this population exhibited a distinctive symptom constellation suggestive of myofascial dysfunction characterized as “persistency*”*: bothersome urinary frequency resulting from bladder discomfort/pelvic pressure conveying a sensation of bladder fullness and a desire to urinate. On examination, 97% of persistency patients demonstrated pelvic floor hypertonicity with either global tenderness or myofascial trigger points, and 92% displayed evidence of impaired muscular relaxation, hallmarks of myofascial dysfunction. We therefore classified this symptom complex “myofascial frequency syndrome”. To confirm this symptom pattern was attributable to the pelvic floor, we confirmed the presence of “persistency” in 68 patients established to have pelvic floor myofascial dysfunction through comprehensive evaluation corroborated by symptom improvement with pelvic floor myofascial release. These symptoms distinguish subjects with myofascial dysfunction from subjects with OAB, IC/BPS, and asymptomatic controls, confirming that myofascial frequency syndrome is a distinct LUTS symptom complex.

**Interpretation:** This study describes a novel, distinct phenotype of LUTS we classified as *myofascial frequency syndrome* in approximately one-third of individuals with urinary frequency. Common symptomatic features encompass elements in other urinary syndromes, such as bladder discomfort, urinary frequency and urge, pelvic pressure, and a sensation of incomplete emptying, causing significant diagnostic confusion for providers. Inadequate recognition of myofascial frequency syndrome may partially explain suboptimal overall treatment outcomes for women with LUTS. Recognition of the distinct symptom features of MFS (persistency) should prompt referral to pelvic floor physical therapy. To improve our understanding and management of this as-yet understudied condition, future studies will need to develop consensus diagnostic criteria and objective tools to assess pelvic floor muscle fitness, ultimately leading to corresponding diagnostic codes.

**Funding:** This work was supported by the AUGS/Duke UrogynCREST Program (R25HD094667 (NICHD)) and by NIDDK K08 DK118176 and Department of Defense PRMRP PR200027, and NIA R03 AG067993.

## Introduction

Storage lower urinary tract symptoms (LUTS), such as urinary frequency, urgency, and bladder discomfort, dramatically impact both urologic well-being and global quality of life. During their lifetime, the majority of women will experience at least one of these symptoms,^1^ which are often chronic and degrade both physical and social functioning.^2^ While many options exist for LUTS management, in reality, clinical care remains inadequate and most women do not seek out care for these symptoms.^3^ For those who attempt to get treatment, many will not receive an accurate diagnosis,^4^ with more than 90% abandoning medical therapies within a year of seeking care.^5^

Accurate diagnosis remains a challenge for defining appropriate management of LUTS. Patients with LUTS are commonly classified into one of several symptom clusters, such as overactive bladder (OAB) and interstitial cystitis/bladder pain syndrome (IC/BPS), with obscure relationships to the pathophysiology underlying those symptoms. OAB is typically distinguished by a sudden urge to urinate with little warning (“urgency”), while interstitial cystitis/bladder pain syndrome (IC/BPS) is distinguished by bladder pain. Yet lacking any definitive diagnostic tests, laboratory markers, or a better understanding of the underlying pathophysiology, these two syndromes are delineated solely by a collection of patient-reported symptoms, making diagnosis subjective and clinician-dependent.^6, 7^ As there is substantial symptomatic overlap between OAB and IC/BPS,^8–10^ we previously proposed a novel nomogram using patient-reported symptoms that has substantial power to distinguish IC/BPS from OAB based on widely used, validated surveys.^11^

Many patients, however, do not fall within the categories of OAB or IC/BPS due to their lack of key features associated with these diagnoses. Many patients present with a similar urinary frequency and desire to urinate (“urge”) as that expressed by OAB patients, but lack the sudden urge to urinate with little warning (“urgency”) that is central to an OAB diagnosis. These patients describe bladder pressure or discomfort, but in the absence of the distinct bladder pain that suggests a diagnosis of IC/BPS. Such patients have been allocated into various categories (e.g., frequency-urgency syndrome), but currently are often labeled as “OAB-dry” to specify this lack of incontinence (or fear of incontinence). While the name suggests a similar spectrum as OAB, the absence of true urinary urgency suggests a different pathophysiology. A recent study of the Lower Urinary Tract Dysfunction Research Network (LURN) cohort revealed that OAB-dry individuals exhibit unique patterns of presenting features that differ from classic OAB subjects, including straining with urination, bladder discomfort, and a feeling of incomplete emptying.^12^ The severity of symptomatic bother in these OAB-dry subjects was most closely related to the severity of pelvic floor tenderness on exam, implicating a possible myofascial dysfunction in this LUTS subgroup.

In the application of our previously described, diagnostic nomogram to a real-world population of subjects with storage LUTS, we hypothesized that the nomogram would successfully identify and classifying subjects with OAB and IC/BPS, but also identify patient subgroups outside the classical LUTS diagnostic paradigm. This analysis describes the features of that group of highly bothered patients who lacked the bladder pain or urgency incontinence characteristic of IC/BPS or OAB, but were symptomatic with urinary frequency, an uncomfortable urge to void without incontinence and bladder pressure, similar to the OAB-dry subset described within the LURN cohort.^12^ In further characterizing this inadequately described phenotype of LUTS, we sought to confirm previously suggested associations with pelvic floor myofascial dysfunction, defining a novel subtype of storage LUTS we termed myofascial frequency syndrome (MFS).

## Materials and Methods

### Study Inclusion

This study was approved by the local Institutional Review Board (IRB#00040261). Female subjects seeking care from urologists board-certified in female pelvic medicine and reconstructive surgery (FPMRS) were administered four validated, written questionnaires at their initial clinical evaluation: 1) the female Genitourinary Pain Index (fGUPI),^13^ 2) Overactive Bladder Questionnaire (OAB-q),^14^ 3) Pelvic Floor Distress Inventory (PFDI-20),^15^ and 4) the O’Leary−Sant Indices.^16^ The fGUPI measures the nature and severity of genitourinary pain, and contains subscales assessing pain, urinary symptoms, and quality of life ^13^. The OAB-q measures both continent and incontinent OAB symptoms and contains subscales for symptom bother, coping behaviors, concern/worry, social interaction, sleep, and health-related quality of life.^14^ Only the symptom questions (1-8) were utilized in this analysis; the 25 health-related quality of life questions on the OABq (OABq 9-33), which address the impact of symptoms on activities and quality of life, were not examined for symptomatic discrimination. The PFDI-20 measures pelvic floor symptoms and includes domains that assess urinary [Urinary Distress Inventory (UDI-6)], defecatory [Colorectal-Anal Distress Inventory (CRADI-8)], and prolapse symptoms [Pelvic Organ Prolapse Distress Inventory (POPDI-6)].^15^ The O’Leary−Sant Indices, which include the Interstitial Cystitis Symptom and Problem Indices (ICSI/ICPI), are commonly used together to measure the severity of and bother associated with urinary frequency, urgency, nocturia, and bladder pain.^16^

### Study Cohorts

Two distinct groups of subjects were analyzed, an *Exploratory cohort* and a *Reassessment cohort* (**Fig. 1**). Patients who were not significantly bothered by their symptoms (i.e., Bother Index <5) were excluded from analysis. Retrospective assessment of the diagnoses coded for these excluded subjects revealed that most sought care for conditions such as asymptomatic microscopic hematuria, nephrolithiasis, or pyuria. Patients with active or recurrent symptomatic urinary tract infections (UTI), prior invasive urinary tract therapies (such as neuromodulation, onabotulinumtoxinA injection, transurethral resection of the bladder, etc.), active smoking, current pregnancy, diabetes, or neurogenic lower urinary tract dysfunction (such as from spinal cord injury, multiple sclerosis, or spinal dysraphism) were excluded. Patients with cyclic pain at menses with concern for endometriosis were also excluded; however, patients with other comorbid functional pain syndromes, such as irritable bowel syndrome or fibromyalgia, were allowed to participate.

**Figure 1.**
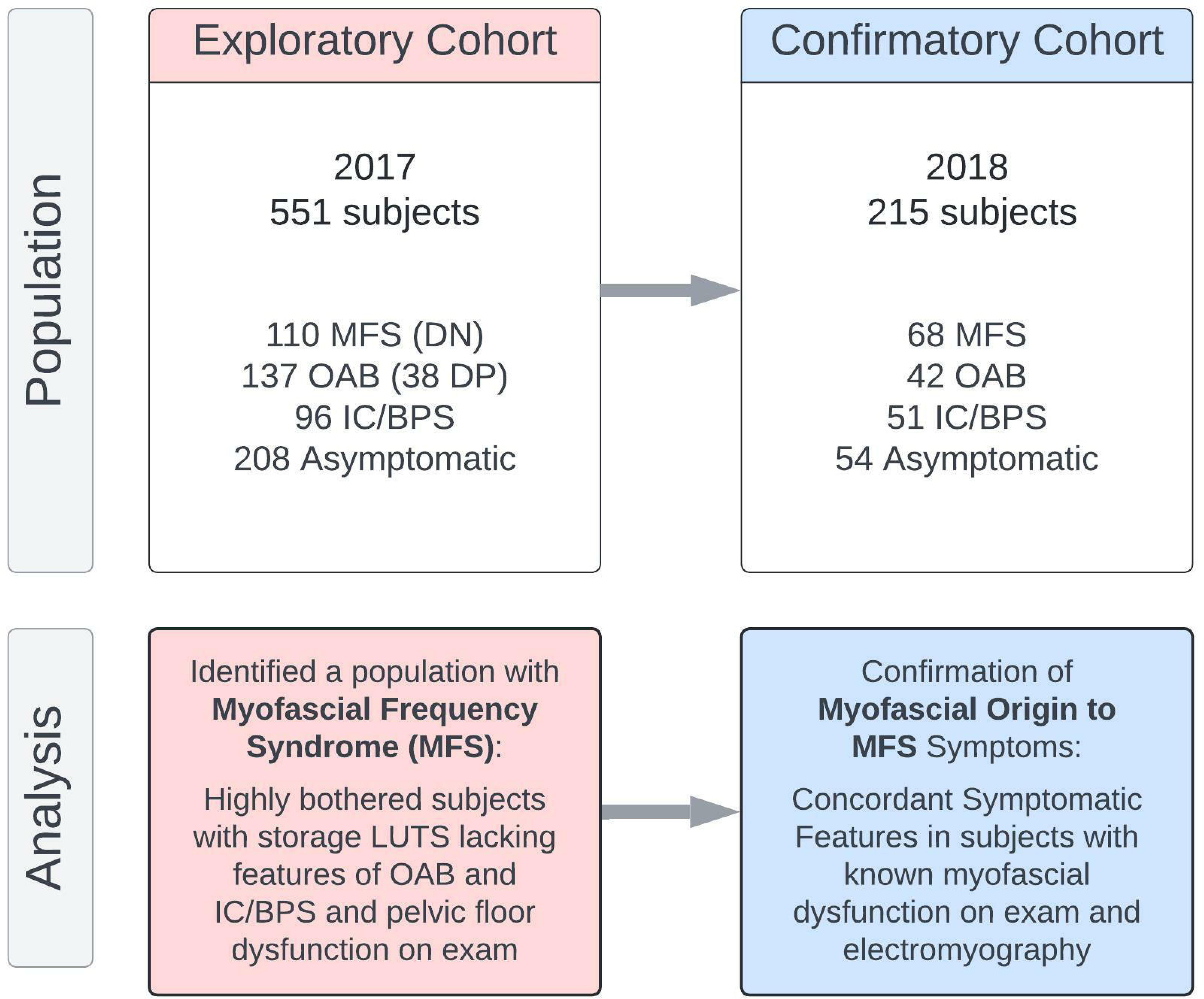
***Study Design.*** The *Exploratory cohort* served initially to investigate what symptomatic complaints and exam findings were common to patients presenting with bothersome LUTS who did not exhibit the classic symptoms of overactive bladder (OAB) or interstitial cystitis/bladder pain syndrome (IC/BPS). After identifying that these patients displayed multiple features of myofascial dysfunction, a phenotype we dubbed “myofascial frequency syndrome” (MFS), we verified the most salient features of MFS by comparison of a group of subjects with LUTS, abnormal pelvic floor findings on exam and EMG findings of a tonically contracted pelvic floor to asymptomatic subjects and patients with confirmed OAB and IC/BPS (*Reassessment cohort*).

The *Exploratory cohort* consisted of 551 consecutive female subjects with a range of irritative storage LUTS evaluated at a tertiary urology practice between January and December 2017. The previously described diagnostic nomogram was applied to this population to classify subjects with characteristic urgency incontinence (UI) as OAB (UICI > 2.5, BPCI < 3), subjects reporting bladder pain (BP) as IC/BPS (BPCI >3, UICI <2.5), and subjects with low bother scores (Bother Index ≤ 5) as controls.^11^ A mixed group of 110 patients were unclassified (UICI < 2.5, BPCI <3, Bother > 5), and represented a range of symptomatic diagnoses such as urinary frequency, urgency, and bladder pressure. Validated questionnaire responses were used to characterize the symptomatic patterns distinguishing this group from the other classifications.

A *Reassessment cohort* of 215 subjects evaluated between January and December 2018 was employed to examine the correlation of distinct symptomatic LUTS patterns. This population contained a myofascial dysfunction group of 68 subjects with urinary symptoms and demonstrable myofascial discomfort or trigger points on physical exam confirmed by non-relaxing pelvic floor hypertonicity on electromyography. Myofascial dysfunction was further implicated as the source of their symptomatology by demonstrable improvements in their urinary symptoms after myofascial release-based, pelvic floor physical therapy (PFPT) or biofeedback. This cohort contained two additional groups with LUTS: 1) 42 subjects diagnosed with OAB who endorsed significant urgency incontinence, lacked any bladder pain on exam or questionnaire assessment, and exhibited detrusor overactivity on urodynamic assessment, and 2) 51 subjects with a clinical diagnosis of IC/BPS with significant bladder pain on exam and questionnaires but who lacked incontinence. An additional 54 subjects with asymptomatic scores on all questionnaires served as controls.

### Qualitative Assessment of Bothered Subjects with LUTS Lacking Pain or Urge

For subjects from the *Exploratory cohort* lacking both urgency incontinence and bladder pain [double-negative (DN) for pain and urgency], we captured findings on discriminant pelvic exams from initial consultations, including prolapse grade by compartment, the presence or absence of stress incontinence on cough stress test, pelvic floor hypertonicity, myofascial trigger points, Oxford grade for pelvic floor muscle strength, and the presence or absence of appropriate relaxation after attempted Kegel.

To determine the prevalent symptomatic features common to this population, we additionally performed a modified thematic analysis.^17^ Descriptions of patient complaints and features were captured from the history of present illness and assessment in the initial consultation within the electronic medical record. Primary urinary complaints, additional bothersome symptoms, aggravating and relieving factors, compensatory behaviors, and terminology used to describe their urinary symptoms were catalogued by two reviewers into primary codes. These primary codes (263 in total) were compiled into common codes capturing similar concepts by each reviewer. A consensus was reached between reviewers regarding conceptual accuracy, resulting in 40 common codes across the population. These codes were clustered into 16 subcategories. The subcategories clustered into 7 categories, which were combined into 3 major themes. In addition, the diagnostic codes associated with both the specialist referral and consultation encounter were recorded.

### Statistical analysis

Bivariate differences between phenotypic groups were examined in the *Exploratory cohort* using Welch’s t-test and chi-square tests. In the *Reassessment cohort,* bivariate logistic regression models predicting MFS were used to examine the magnitude of the odds ratios for individual symptomatic features. Measures with significant, positive associations with MFS were included in a multivariable model predicting MFS. All analyses were conducted in Stata version 16.1, Stata Corp (College Station, Texas, USA).

## Results

### A highly bothered subset of patients lacks clear bladder pain or urgency/urge incontinence

The *Exploratory cohort* included 551 female subjects sequentially presenting to a tertiary urology clinic in the 2017 calendar year (**Fig. 1**). Of these, 208 subjects were not significantly bothered by genitourinary complaints (Bother Index < 5), leaving 343 patients dissatisfied with their urinary symptoms. Application of the previously described storage LUTS nomogram (**Fig. 2**) classified 96 (28%) subjects as IC/BPS, defined by a bladder pain composite index (BPCI) score ≥ 3 in the absence of significant urinary urgency. This subgroup exhibited symptoms consistent with that diagnosis, demonstrating the highest levels of bladder pain (ICSI4, ICPI4) aggravated by filling (fGUPI 2c, 2d) (**Table 1**). An additional 137 subjects (40%) were categorized as OAB [urge incontinence composite index (UICI) scores ≥ 2.5], with characteristically high scores describing urgency associated with a fear of incontinence (UDI-6 2, OABq8, ICSI1, ICPI3; **Table 1**). Within the OAB group, there were 38 (11%) subjects with urgency and urgency-related incontinence as their dominant symptoms, but who also had substantial bladder pain (BPCI ≥ 3) conveyed on questionnaires [double-positive (DP) for pain and urgency].

**Figure 2.**
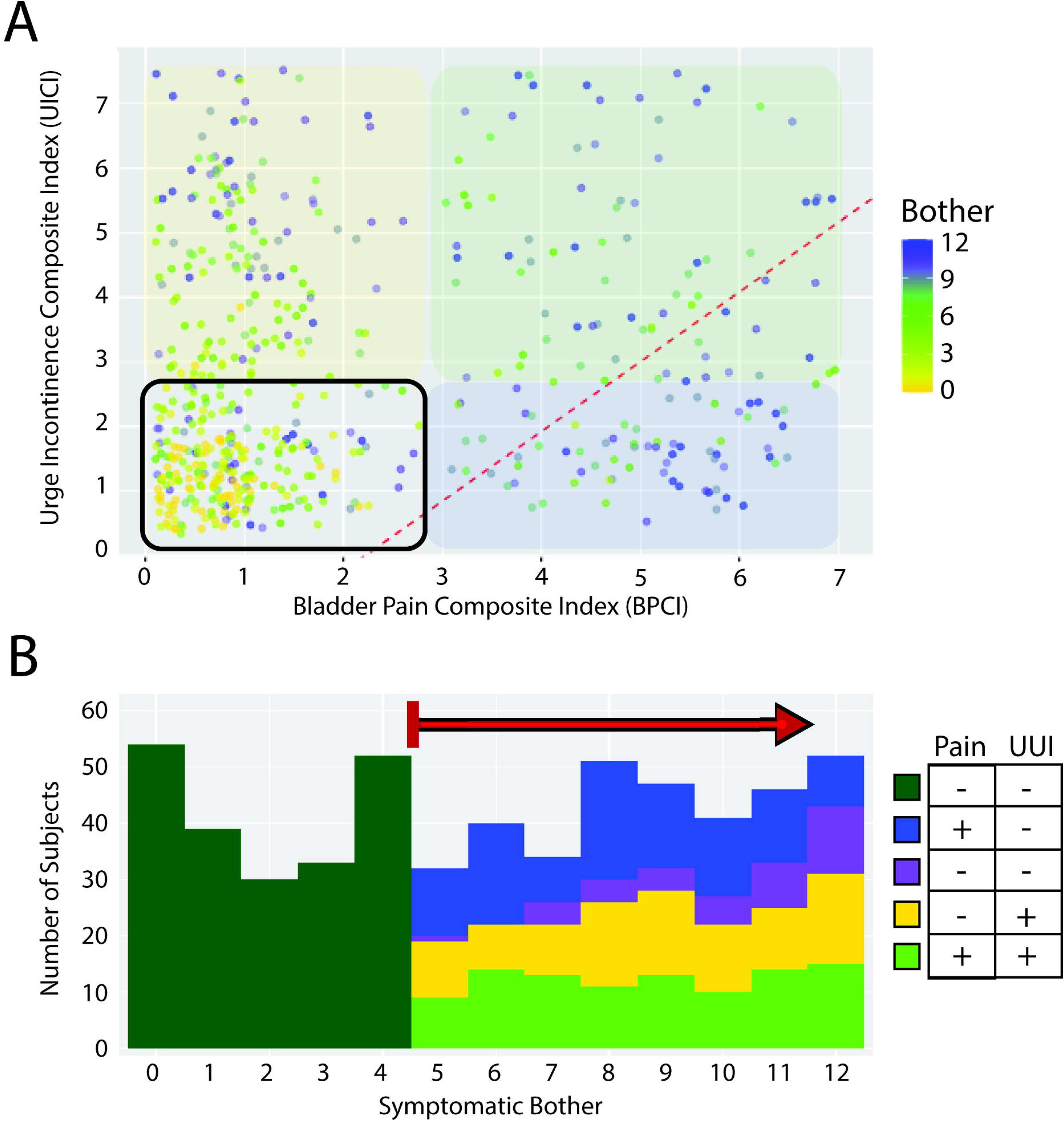
***A highly bothered subset of patients lacks clear bladder pain or urgency/urge incontinence.*** A) In a test population of 551 subjects with a variety of LUTS, the Bother Index identified those patients dissatisfied with their urinary symptoms (yellow to blue heat map). B) A dot plot of BPCI and UICI symptomatic scores for all subjects with high bother (green gradient indicating bother is as in A) displays the degrees of bladder pain (BP) and urgency/urge incontinence (UI), respectively. OAB patients with UI (yellow overlay) occupy the upper left quadrant, while IC/BPS patients with BP (blue overlay) are in the lower left quadrant. A substantial population negative for the symptom domains originally described for this diagnostic LUTS nomogram is seen in the left lower quadrant, dubbed the Double Negative (DN) group (purple). These data also reveal a population with both UI and BP in the upper right quadrant (green; double Positive: DP). The division previously defined between OAB and IC/BPS patients from our prior nomogram is indicated by the red dotted line.

**Figure 3.**
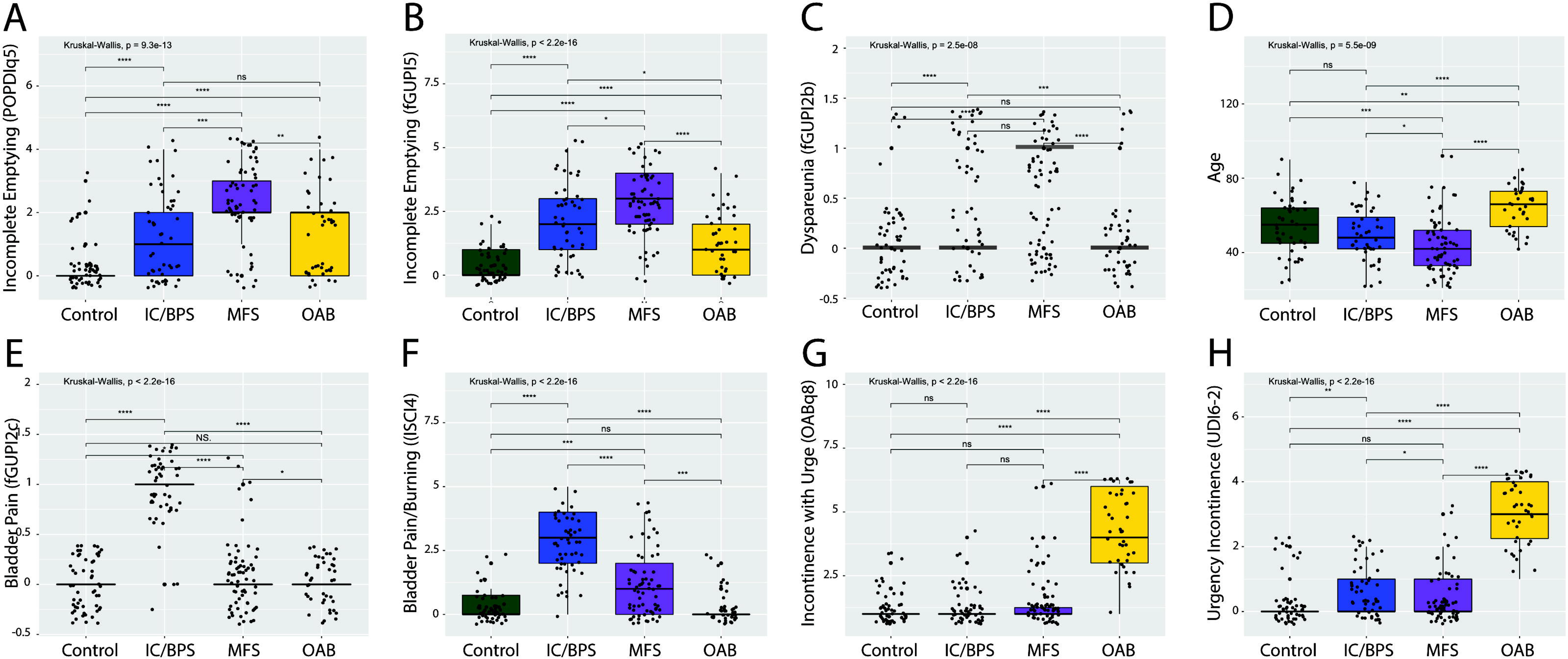
***In the* Reassessment cohort*, MFS subjects exhibit symptomatic features distinct from OAB and IC/BPS subjects.*** While multiple symptomatic features, such as urinary frequency, were increased in all LUTS groups, the MFS demonstrated significantly increased severity of a sensation (A) and frequency (fB) of incomplete emptying and dyspareunia (C). Subjects with MFS were also significant younger than the other populations (D). In contrast, subjects with IC/BPS exhibited higher levels of pain with bladder filling (E) and bladder pain and burning (F), while OAB subjects demonstrated consistent elevations in the sudden incontinence associated with urgency (G) and a strong desire to urinate (H). Brackets indicate pairwise comparisons by Wilcoxon rank sum tests. * p<0.05, ** p<0.005, *** P<0.005, ns: not significant.

**Figure 4.**
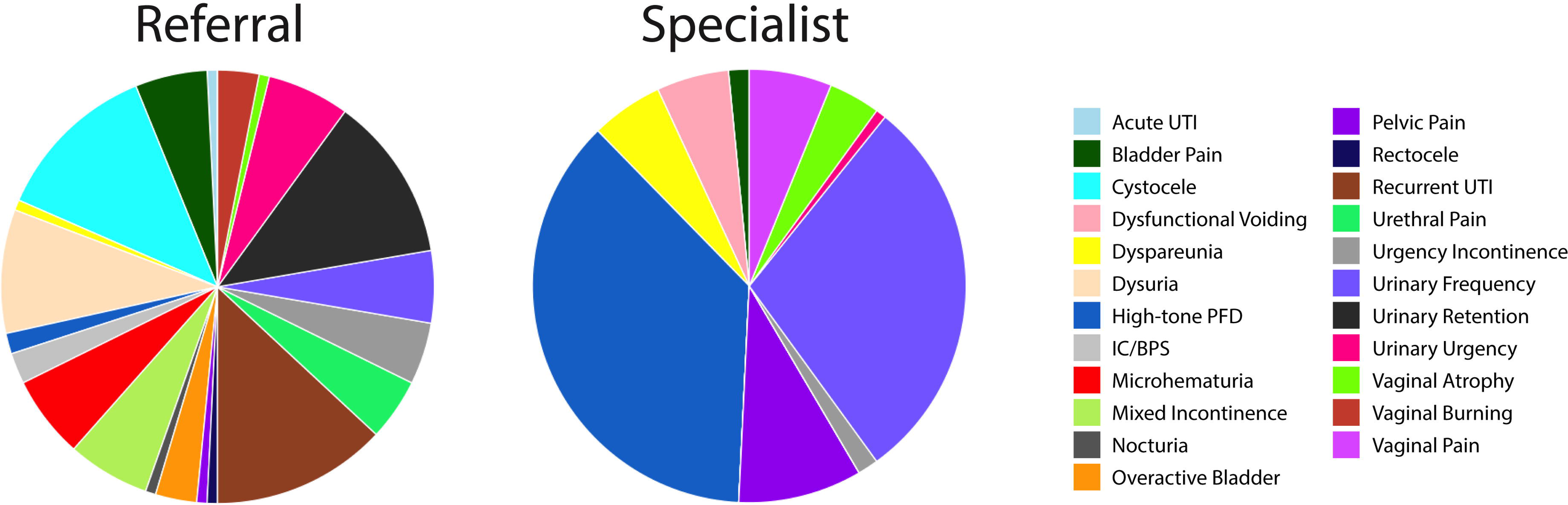
***Referral and Specialist Diagnosis for Myofascial Frequency Syndrome Subjects in the* Reassessment cohort.** At referral, subjects with myofascial frequency syndrome were frequently diagnosed with a wide range of presumptive diagnoses. As there is no specific code for LUTS derived from a myofascial dysfunction, the coded diagnoses reflected the typical symptoms of this disorder (e.g., urinary frequency, pelvic pain, vaginal pain, dyspareunia) and pelvic floor muscle dysfunction (high-tone pelvic floor dysfunction, dysfunctional voiding).

**Table 1.**
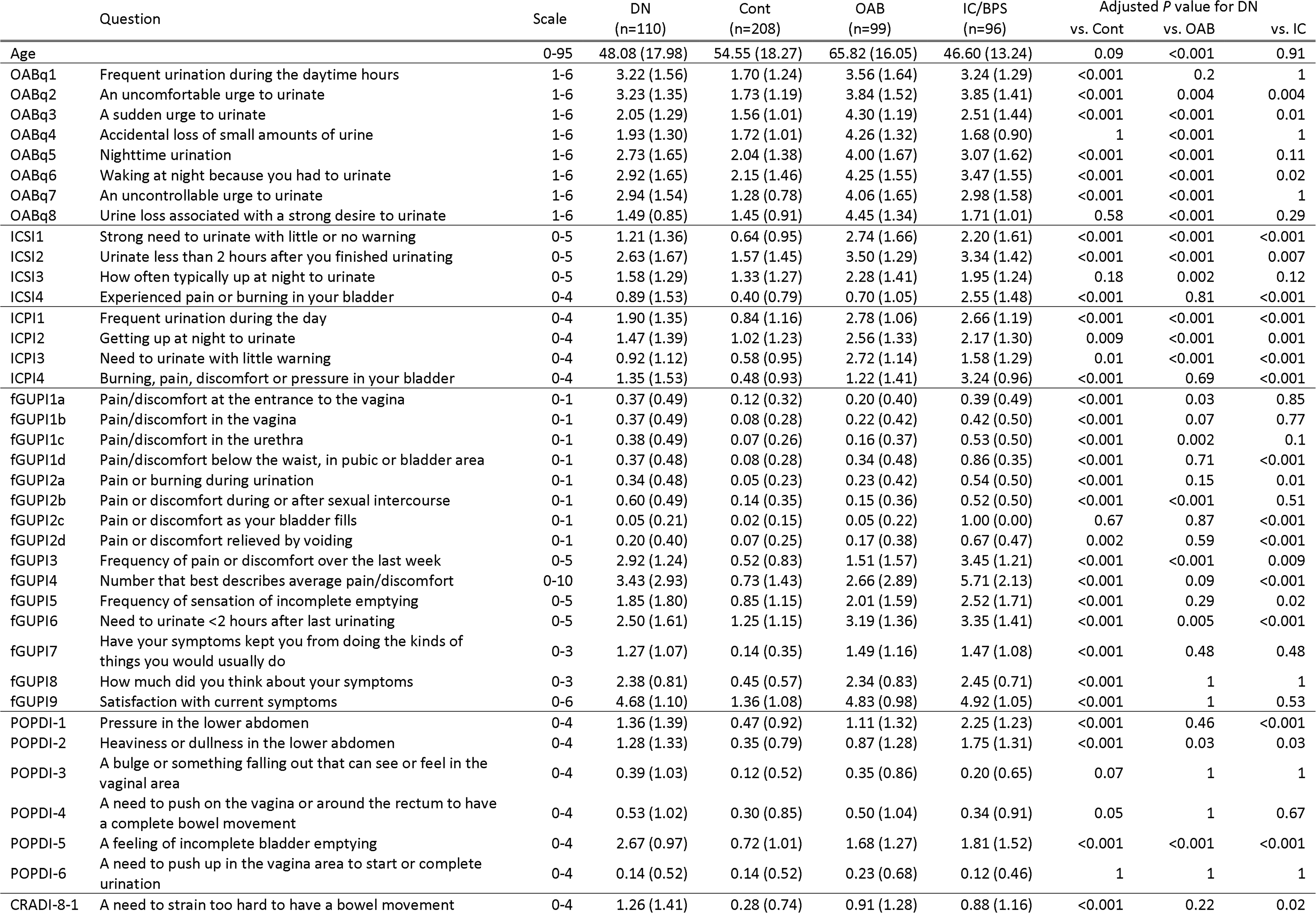
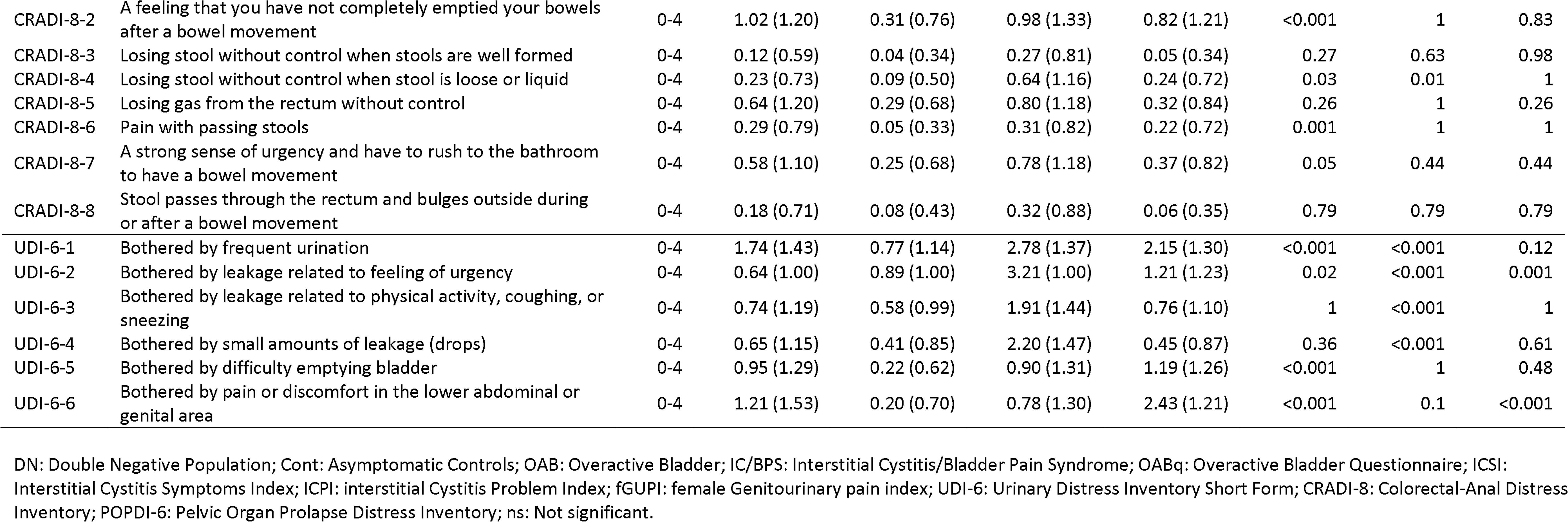
Symptomatic Features of the Exploratory cohort by Nomogram Quadrant.

The remaining 110 patients (32%) demonstrated significantly bothersome urinary symptoms but lacked either the bladder pain or urgency with fear of incontinence that characterized the IC/BPS and OAB groups, respectively (**Supplemental Fig. 1**). Despite lacking pain or urge incontinence, this DN population was bothered at similar levels to the other subject groups and was younger than the other populations.

### The DN population exhibits a symptomatic profile distinct from other LUTS symptom clusters

To identify the nature and severity of bothersome genitourinary symptoms present in the DN population, we examined patient-reported scores on 51 individual questions from validated instruments (**Table 1**). Individual questions were clustered into redundant groups to attempt to identify specific symptoms common to and shared by this DN population. Only two symptomatic features demonstrated higher values in this population than in the other groups: *a sensation of incomplete bladder emptying* (POPDI-6 5) and *a need to strain with defecation* (CRADI-8 1). While these *sensations* of incomplete bladder and bowel emptying were significant in comparison to controls, companion questions describing the actual “difficulty emptying the bladder” (UDI-6 5) or “difficulty emptying your bowels at the end of a bowel movement” (CRADI-8 2) were not significantly different from other LUTS populations.

In addition, the endorsement of urgency differed based on the individual wording of the questionnaire (**Table 1**): DN subjects endorsed a similar magnitude of urge as classic OAB patients when expressed as a “strong” or “uncomfortable” need to urinate (OABq2). However, this population was minimally symptomatic and significantly different from OAB subjects when the urge was expressed as a “need to urinate with little warning” (ICSI1, ICPI2) or “sudden urge” (OABq3).

A similar pattern was seen for pain symptoms, with the questions divided into two categories. Scores for the DN population were similar to the IC/BPS population when the question wording included the terms “pressure” or “discomfort” alone or in combination with other pain terms (ICPI4) but were significantly less when the terms “burning” or “pain” were used in isolation (ICSI4). The patterns of pain localization also differed between the DN and IC/BPS populations. IC/BPS subjects were characterized primarily by pain with bladder filling relieved by bladder emptying (fGUPI 2c, 2d). In contrast, the DN population exhibited additional, variable manifestations of genitourinary pain, but only pain related to intercourse (fGUPI 2b) was present in more than half of DN subjects (**Table 1**).

Urinary frequency was a dominant aspect of this DN population (OABq1, ICSI2, ICPI3, UDI-6 1), although the objective severity and bother attributed to this symptom was lower than that seen for OAB or IC/BPS subjects (**Table 1**). Urinary incontinence (OABq4, OABq8, UDI-6 1-3), fecal urgency/incontinence (CRADI-8 3-5, CRADI-8 7), and nocturia (OABq5, ICSI3, ICPI2) were low in this population, clearly distinguishing this group from the classic OAB population. Thus, the dominant features of the DN population in validated questionnaires were a perception of incomplete emptying, urinary frequency, pelvic discomfort, pelvic pressure unrelated to bladder filling, and straining to void/defecate without sudden-onset urgency, nocturia, significant incontinence, or bladder-specific pain.

### The double-negative population exhibits multiple aspects of myofascial dysfunction

As symptom scores on the Colorectal-Anal Distress Inventory (CRADI-8) were similar in pattern to patients with pelvic floor dyssynergia described in the colorectal literature^18^, we examined the diagnoses coded in association with the initial specialist consultation of the 110 subjects classified into the DN group. Ninety-seven (88%) of these subjects carried a coded diagnosis of high-tone pelvic floor dysfunction at initial evaluation. The remainder of the patient visits carried codes associated with symptomatic features, such as urinary frequency, dysuria, and pelvic pain.

Eighty-four (76%) of DN subjects had positive documentation of increased pelvic floor hypertonicity with either global tenderness or distinct, painful myofascial trigger points, despite pain not being a primary complaint of the patient without elicitation. Only 3 (3%) of the subjects in this group did not have any evidence of myofascial hypertonicity or tenderness documented on physical exam; most subjects with sufficient assessment (67/73, 92% of those assessed, 61% overall) also had evidence of impaired muscular relaxation after voluntary pelvic floor contraction, a hallmark of myofascial dysfunction. We therefore named the symptom complex experienced by the DN population “myofascial frequency syndrome” to reflect both LUTS and myofascial dysfunction.

### Thematic analysis of DN patient histories links uncomfortable bladder pressure to urinary frequency

Qualitative review of individual histories of present illness using thematic analysis (**Table 2**) confirmed the pattern of symptomatology revealed by the questionnaires, identifying bothersome urinary frequency resulting from a perception of bladder pressure or discomfort and conveying the sensation of fullness with an uncomfortable need to urinate as common among these patients, a symptomatic constellation we termed “persistency”. Several additional symptomatic features that could not be captured by questionnaires were common among these subjects, such as a minimal or positive effect of fluid intake on symptom severity, minimal nocturia once asleep with increases in frequent urination experienced with initially lying down at night, worsening of symptoms with certain activities (e.g., travel, extended periods of sitting, or very intense exercise), a sensation of bladder fullness unrelieved by voiding, and occasional insensate, small volume incontinence. These highly bothered subjects often had no clear dominant symptom, but almost all complained of “persistency”: urinary frequency resulting from an uncomfortable inability to defer voiding combined with a sensation of incomplete emptying and pelvic discomfort that was difficult to localize.

**Table 2:** Thematic Analysis of Presenting Features of Patients with MFS. Descriptions of unique symptomatic features were extracted from the history of present illness (HPI) at each patient’s initial consultation.

| Global Theme | Category | Subcategory | Code | Raw data (examples) |
| --- | --- | --- | --- | --- |
| Urinary frequency | Persistent need to void | Constant frequency | Persistent feeling of needing to void | "She can never be comfortable for any period of time, even right after voiding" |
|  |  |  | Fullness out of proportion with volume | "She states that there is never very much there, but it feels like I'm going to burst, so I have to go." |
|  |  |  | Intrusive urge | "She notes the need to go is powerful, and very hard to ignore." |
|  |  | Prone frequency without nocturia | Nighttime Frequency | "She has to go to the bathroom 5-6 times before she can finally get to sleep" |
|  |  |  | Association with lying down | "Very little comes out each time, but 5 minutes after she gets back in bed, she feels like she has to go again to get empty." |
|  |  |  | No Nocturia | "Once she gets to sleep, she stays asleep for ~6 hours, but has to go multiple times when she first lies down." |
|  | Activities exacerbate symptoms | Excessive physical activity | Exercise brings on urge | "She cannot exercise without feeling like she has to urinate every 20 minutes" |
|  |  |  | Physical activity worsens symptoms | "The day after a lot of walking, like a trip to Disneyland, will be much worse" |
|  |  | Immobility for long periods of time | Worsened by immobility | "Travelling in the car for long trips is terrifying because it will make these symptoms so much worse." |
|  |  |  | Symptoms persist after immobility | "Always gets the feeling of needing to go constantly right after a long plane flight, almost like the beginning of a UTI but it never progresses." |
|  |  | Extended Periods of Sitting | Sitting worse than other positions | "She has to lie down in the back seat on long car trips, or she will have to urinate every 10 minutes." |
|  |  |  | Sitting increases urge | "She notes that she dreads long meetings, cause the urgency is so much worse sitting in one place for that long and she feels like she can't go or her coworkers will think something is wrong with her." |
| Sensation of Bladder Fullness | Sensation of incomplete emptying | Constant sensation of fullness | Persistent urge | "it always feels like there is something there, even right after going" |
|  |  |  | Rapid return of fullness | "Within 15 minutes of going, she feels like her bladder is completely full again." |
|  |  |  | Perception of fullness | "She states she knows there's nothing there, but feels like she is still full." |
|  |  | Poor Toileting Behaviors | Straining to Void | "She sits on the toilet for at least a few minutes after trying to push any remaining urine out, even though little comes, just to make sure she is empty" |
|  |  |  | Double Voiding | "She will typically get up and then go right back to the toilet and only a little bit more will come out." |
|  |  |  | Persistent Voiding | "She complains that she is losing a lot of time to just sitting on the toilet in an attempt to finally feel empty" |
|  | Bladder discomfort and Pressure | Pressure | Building pressure | "It is not a sudden-onset urgency, more of a building pressure that is hard to ignore." |
|  |  |  | Pressure throughout pelvis | "there is a pressure that begins midline over the bladder and then spreads across the low pelvis if she tries to hold it." |
|  |  |  | Heaviness | "there is a heaviness in the pelvis that is always there, it just gets worse the longer she tried to wait before urinating." |
|  |  | Discomfort | Discomfort builds | "If she does not go when the desire comes, she will not be incontinent, but will be more and more uncomfortable until she just has to go" |
|  |  |  | Pain with deferring urination | "If she had to hold it past the early sensation of needing to go, it would just get painful to hold it, so she never does." |
|  |  |  | Uncomfortable urge | "it is not painful, but just a very uncomfortable pressure that interferes with her ability to do anything else." |
| Uncomfortable Urge, not Urgency | Minimal Impact of Fluids | Little dependence on fluid | No impact of fluid limitation | "if she drinks more, she goes more, but even when she limits fluids heavily, the feeling of constantly needing to void never goes away." |
|  |  |  | Low volume urge | "She states as soon as she takes a sip of any fluids, the urine hits her bladder and she has to go again" |
|  |  |  | Frequency independent of intake | "It doesn't matter how little she drinks, the feeling [of bladder fullness] never goes away." |
|  |  | Improved voiding with more volume | better voiding efficiency with more fluids | "she notes that if she drinks more, sometimes it will help her feel like she can empty better and be more comfortable" |
|  |  |  | no impairment in capacity | "She notes that the only time she feels she can really get empty is first thing in the morning when her bladder is really full." |
|  | Insensate Incontinence | Activity-associated leakage | Worse leak with intense activity | "While exercising, never feels any leakage, but notices that she can be very wet afterwards" |
|  |  |  | Minor activities bring on leakage | "Never feels a gush, but notes her liner is pretty wet when she gets home after a long walk." |
|  |  | No awareness of leaks | Feeling of wetness, not leak | "she never notices anything coming out, but when she gets undressed at the end of the day, she notices her underwear is wet." |
|  |  |  | Constant incontinence | "No particular events or activities bring it on, she just notices a little wetness in her underwear when she goes to the bathroom." |
|  |  | Not stress incontinence | No leakage with stress maneuvers | "denies any leakage with cough, laugh or sneeze, but feels a little bit wet all throughout the day" |
|  |  |  | No association with fullness | "She denies any period when she can be predictably dry for any length of time" |
|  |  | Very low volume | Bothered by smell | "never soaks through the inner cotton layer of her underwear, but definitely smells like urine, so she keeps a change of underwear in her purse." |
|  |  |  | Low volume | "she just feels a little bit wet, all throughout the day, but never enough to wear a pad." |
|  | No Urgency Incontinence | No Urgency or Urgency Incontinence | No sudden-onset urgency | "She never leaks or has to run suddenly to the bathroom, but constantly needs to go." |
|  |  |  | No incontinence with urge | "She states if she had to hold it and couldn't get to a bathroom, she could make it." |
|  |  |  | Volitional voiding to avoid discomfort | "She could wait to void, but doesn't want to." |

### Defining the constellation of urinary symptoms associated with pelvic floor myofascial dysfunction

We next sought to confirm the presence of *persistency* in the *Reassessment* cohort, an independent cohort containing patients with a known myofascial origin to their urinary symptoms. From all patients with primary assessment in 2018, we identified 68 patients established through comprehensive diagnostic evaluation to have pelvic floor myofascial dysfunction as the source of their urinary symptoms. Urodynamic testing with pelvic floor electromyography demonstrated a tonically contracted, non-relaxing pelvic floor, even during voiding. Subjective symptomatic improvement in LUTS following pelvic floor physical therapy or biofeedback served as secondary confirmation of myofascial dysfunction as the source of their symptoms. Consistent with the clinical profiles of the MFS group, these individuals had sought care for urinary complaints, typically urinary frequency and a persistent urge to void, in the absence of significant bladder or pelvic pain. These subjects were then combined with 42 subjects with OAB, 51 subjects with IC/BPS, and 54 asymptomatic subjects seen during the same interval to create a *Reassessment cohort*.

Bivariate logistic regression models were used to identify individual patient-reported symptomatic features associated with myofascial dysfunction from the OABq symptom assessment, PFDI-20, fGUPI, and ICSI/ICPI (51 items in total). Nineteen symptom features found to have significant associations were included in multivariable models predicting myofascial dysfunction (**Table 3**), one including and one excluding the asymptomatic controls, to ensure that the identified features could distinguish myofascial dysfunction subjects from both controls and other subjects with genitourinary symptoms. Only two variables remained positively associated with myofascial dysfunction in both models (fGUPI 5, POPDI-6 5), both of which describe a sensation of incomplete bladder emptying. The sensation of lower abdominal pressure (POPDI-6 1) as well as pain with sexual intercourse (fGUPI 2b) were both positively associated with myofascial dysfunction, although the association was not statistically significant in the LUTS-only model (*p* = 0.057). Both urine loss associated with a strong desire to urinate (OABq8) and pain with bladder filling (fGUPI 2c) demonstrated significant negative associations with myofascial dysfunction, clearly distinguishing this group from the OAB and IC/BPS groups, respectively. Thus, this independent cohort of patients with known myofascial dysfunction demonstrated the symptoms of *persistency*, embodied by the sensation of incomplete emptying associated with bladder pressure and dyspareunia, but without urgency, urge incontinence, or pain with bladder filling.

**Table 3.**
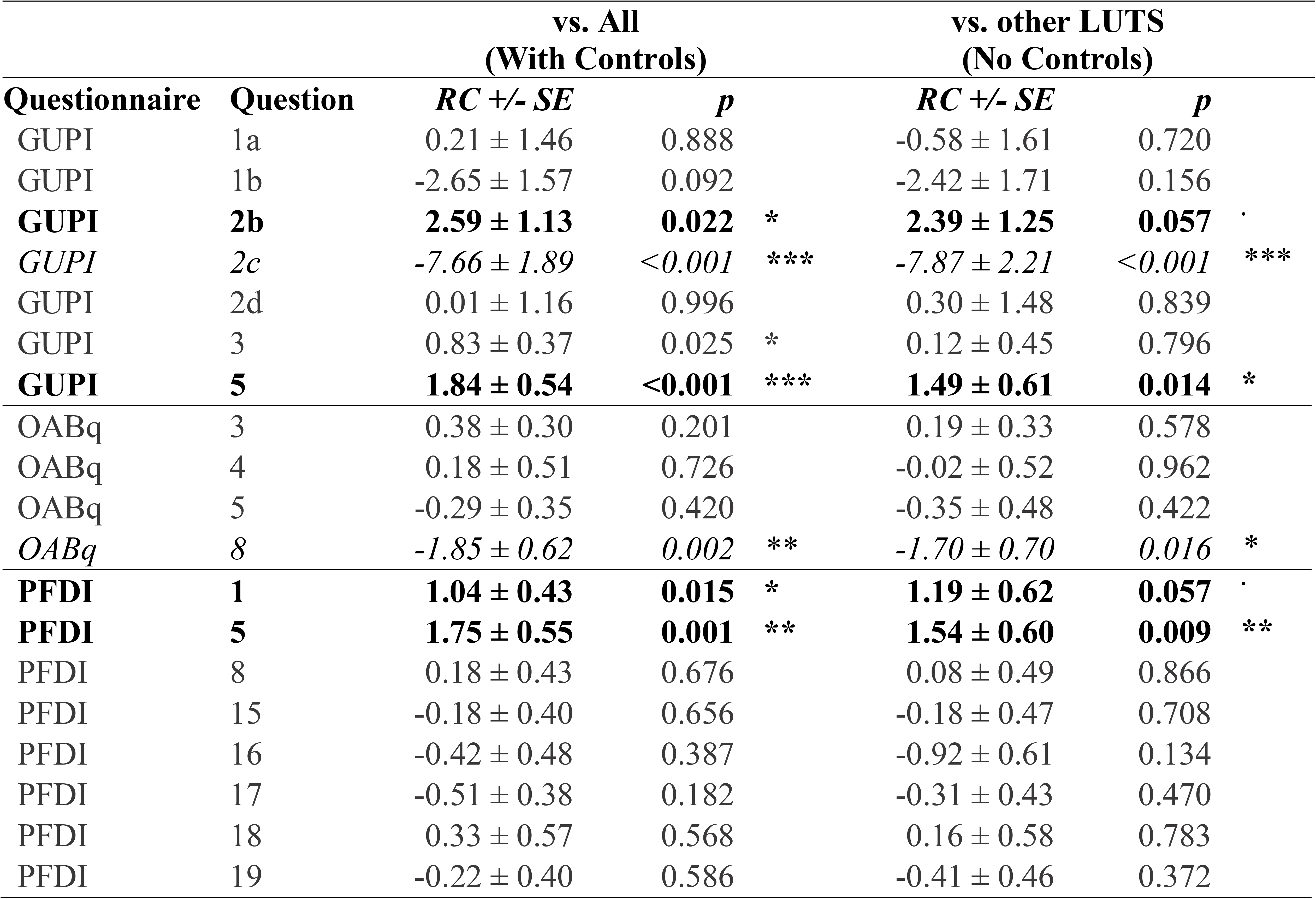
Multivariable Logistic Regression of Symptomatic Features Predicting Myofascial Dysfunction.

### The clinical presentation of DN subjects is associated with significant diagnostic confusion

Pre-referral diagnoses for the myofascial dysfunction patients were highly varied, including cystocele, urinary retention, dysuria, recurrent UTI, hematuria, urethral pain, urinary frequency, and urgency, without a single dominant diagnosis (**Fig. 5A**). After initial specialist evaluation, more than 75% of MFS subjects were given a primary diagnosis of either high-tone pelvic floor dysfunction (37%), urinary frequency (29%) or pelvic pain (9%) (**Fig. 5B**). Other less common diagnoses included dysfunctional voiding (5%), dyspareunia (5%), and vaginal pain (6%), all of which encompass features common to the MFS symptom cluster.

## Discussion

In this study, we describe a previously underrecognized phenotype of storage LUTS that we named myofascial frequency syndrome (MFS), which is distinct from other common forms of storage LUTS such as OAB and IC/BPS. Affected patients typically seek care for nonspecific symptoms of urinary frequency, urgency, pelvic pressure, or a sensation of incomplete bladder emptying, embodied in the symptomatic concept we identified as *persistency*. Demonstrating objective findings of pelvic floor hypertonicity and trigger points on physical exam, MFS subjects demonstrate symptomatic improvements in these LUTS with myofascial-directed therapies, such as PFPT.^19^ MFS was common in this study, identified in 20% of all new urologic consultations and 32% of those presenting with urinary frequency. While MFS shares many features with myofascial pelvic pain/pelvic floor myalgia (MPP/PFM), the presentation of these patients is distinct in that they do not report pain. Although pain can sometimes be elicited or trigger points identified on pelvic exam, patients’ reasons for seeking care are primarily urinary, which can lead to difficulties suspecting the pelvic floor as an etiology of their symptoms if a detailed pelvic floor exam is not performed. While the co-existence of urinary complaints in patients with MPP/PFM is well-documented,^20^ few reports have detailed the urinary manifestations associated with myofascial dysfunction in the absence of baseline bladder/pelvic pain.^21–23^

In the colorectal literature, however, myofascial dysfunction has been well documented to contribute substantially to defecatory symptoms in the absence of pain.^24^ Multiple phenotypes of myofascial dysfunction, including failed, incomplete, or paradoxical relaxation of the levator muscle complex, contribute to distinct patterns of defecatory symptoms, ranging from obstructive defecation to insensate fecal incontinence to painful defecation. Our data suggest that there is a similar breadth of urinary symptomatology attributable to myofascial dysfunction that extends beyond myofascial pain, of which MFS is one such phenotype.

Our analysis reveals that MFS patients commonly present with a combination of symptoms that overlap substantially with other categories of storage LUTS. The clinical behavior of MFS as a “great pretender”, mimicking many of the features of better characterized conditions, may explain why this syndrome is frequently unrecognized and remains poorly documented in the literature. MFS subjects describe urinary frequency often due to an uncomfortable feeling of needing to urinate; many will use the word “urgency” to describe this sensation, making MFS easily confused with OAB, even though this sensation is distinct from true urgency. Subjects with MFS often complain of pelvic pressure and heaviness and straining to defecate, similar to subjects with pelvic organ prolapse, although typically patients will deny a vaginal bulge or a need to splint to void/defecate. In addition, MFS patients will often complain of a bladder/pelvic discomfort that is not described as overt pain, but, in combination with frequency and urge, could easily be conflated with IC/BPS. Some patients even exhibit a fluctuating urethral/vaginal discomfort or burning and a terminal dysuria that can be mistaken for recurrent urinary tract infections, although the baseline symptoms rarely resolve completely between aggravating episodes. The most discriminatory feature, however, was the feature of persistency: a sensation of incomplete bladder emptying that was unexpectedly distinct from the difficulty emptying the bladder experienced in patients with prolapse.

Given the wide range of referral diagnoses for the population with MFS, it is reasonable to conclude that this mixture of symptoms elicits significant confusion for providers. While a standardized screening examination has been developed for identifying and quantifying internal hip and pelvic floor myofascial pain on palpation,^25^ a diagnostic examination requires formal evaluation by a specially trained pelvic floor physical therapist or rehabilitation medicine specialist. Even for skilled urologists and urogynecologists, making a diagnosis of MFS requires both a careful, discriminant pelvic exam and a detailed symptomatic history to identify potential myofascial dysfunction. This can be challenging in an era in which telemedicine is increasingly an expectation by patients and when the amount of time allowed for visits may limit the ability to perform this assessment adequately. Future derivation of an objective symptomatic measure associated with this phenotype could greatly assist providers in suspecting a myofascial origin to a patient’s urinary symptoms and focusing the physical assessment and possible treatments to be considered.^19^

An additional limitation to improving the diagnosis of MFS is the absence of consensus terminology for myofascial pelvic floor dysfunction. For example, numerous terms have been used to describe a pelvic floor with abnormal pelvic floor contractile activity, including: nonfunctioning pelvic floor muscles, short pelvic floor syndrome, atonic or acontractile pelvic floor, hypertonic pelvic floor, high-tone pelvic floor dysfunction, and pelvic floor/levator muscle spasm.^26^ The International Continence Society^27^ proposed classification of pelvic floor muscle dysfunction into overactive, underactive and non-functioning categories, but these general descriptions fail to capture the range of possible pathologies, noting only their association with genitourinary pain as well as voiding, defecatory, and sexual dysfunction. There is also a lack of diagnostic ICD-10 codes describing this specific pelvic floor condition, further confounding epidemiologic study of myofascial dysfunction. As we learn more about the role of the pelvic floor in urinary symptomatology, it is imperative we develop both consistent terminology and diagnostic codes to reflect these conditions. Common terminology is one necessary and surmountable step to improving the awareness and care for these patients moving forward; its deficiency limits the reproducibility of this analysis as no gold standard for the diagnosis of MFS has yet been defined.

Diagnosis of MFS could be achieved with objective, reproducible, and quantifiable assessments of pelvic floor muscle fitness. Such measures may help differentiate between forms of myofascial dysfunction and correlate those patterns with different symptomatologies, as has been done with defecatory disorders. Unfortunately, there are currently no accepted measures to quantify muscular fitness or dynamic myofascial function for the pelvic floor. While pelvic floor electromyography can identify paradoxical contraction during voiding that is characteristic of dysfunctional voiding, it is not reliable in assessing pelvic floor muscle strength or fitness/functionality.^28^ Novel pelvic floor assessment methods that identify and objectively measure the types of myofascial dysfunction are greatly needed. Urinary symptoms may be caused by a shortened, acontractile pelvic floor, a hypertonic, fixed pelvic floor, or a dyssynergic pelvic floor. The causes, manifestations, and treatments for each of these pelvic floor dysfunctions may be unique; a better understanding of how to prevent and manage each etiology requires more advanced pelvic floor assessment.

The limitations of this analysis bear mention. As this cohort included only women, utilizing multiple symptomatic measures specifics to women, it is not clear how these symptoms present in men and how myofascial dysfunction manifests as a cause for LUTS. In addition, our thematic analysis utilized the provider documentation of patient symptoms. While significant detail in these records was sufficient to gain substantial insight into the experiences of patients, future studies should attempt to capture these experiences directly from the patient perspective.

While MFS can occur in subjects across the age spectrum, it comprised a larger proportion of younger patients presenting with LUTS. One explanation for this pattern is that insults to the pelvic floor that accompany aging (e.g., pelvic floor obstetrical trauma, increased muscular laxity) may be protective against MFS. However, OAB incidence also increases dramatically at the menopausal transition and continues to increase thereafter. Subjects with concurrent urgency incontinence characteristic of OAB would not have been classified as MFS, as this group was defined as lacking this symptom. Therefore, it is unclear but likely that MFS co-exists with other storage LUTS, particularly in older patients. Many patients with IC/BPS demonstrate pelvic floor hypertonicity on exam and benefit significantly from PFPT,^29–31^ suggesting that multiple forms of storage LUTS can coexist in a single patient. Hopefully, as we gain better recognition and tools for the evaluation of MFS, we will be able to begin to dissect out the relative contributions of neurologic, inflammatory, detrusor, and myofascial dysfunction in these more complex clinical presentations.

Regardless of these limitations, this study takes the first step of defining and differentiating MFS, a myofascial-driven, non-painful urologic symptom cluster, as a distinct storage LUTS subtype characterized by “persistency”: an uncomfortable, persistent desire to urinate associated with urinary frequency and a sensation of incomplete bladder emptying.

## Data Availability

All data produced in the present study are available upon reasonable request to the authors for research purposes.

## Supplemental Figure Legends

**Supplemental Figure 1. *Symptomatic Features of the* Exploration cohort.** Scores for overall bother (A), ages (B), urgency incontinence (C; UICI), and bladder pain (D; BPCI) were compared for the highly bothered subjects in each of the four quadrants in comparison to control subjects. Cont: controls; DN: UUI-BP-; OAB: overactive bladder (UUI+); DP: UUI+, BP+; IC/BPS: interstitial cystitis/bladder pain syndrome (BP+). UUI: Urgency Urinary Incontinence. BP Bladder Pain.

## Abbreviations

AUC,: Area Under Curve
BP,: Bladder Pain
BPCI,: Bladder Pain Composite Index
BMI,: Body Mass Index
DN,: Colorectal-Anal Distress Inventory (CRADI-8), Double Negative
DP,: Double Positive
fGUPI,: female Genitourinary Pain Index
FPMRS,: Female Pelvic Medicine and Reconstructive Surgery
ICSI,: Interstitial Cystitis/Bladder Pain Syndrome (IC/BPS), Interstitial Cystitis Symptom Index
ICPI,: Interstitial Cystitis Problem Index
LASSO,: Least Angle Shrinkage and Selection Operator
LUTS,: Lower Urinary Tract Symptoms
MFS,: Myofascial Frequency Syndrome
OAB,: Myofascial Pelvic Pain/Pelvic Floor Myalgia (MPP/PFM), Overactive Bladder
OABq,: Overactive Bladder questionnaire
PCI,: Persistency Composite Index
PFDI,: Pelvic Floor Distress Index – 20
PFPT,: Pelvic Floor Physical Therapy
(POPDI-6),: Pelvic Organ Prolapse Distress Inventory 6
(p-CLUS),: Phenotyping of Comprehensive Lower Urinary Symptoms
PA,: Predictive Accuracy
UI,: Urgency Incontinence
UICI,: Urge Incontinence Composite Index
(UDI-6),: Urinary Distress Inventory 6
(UTI),: Urinary Tract Infection

## Notes

**Funding:** This work was supported by the AUGS/Duke UrogynCREST (Urogynecology Clinical Research Educational Scientist Training) Program (R25HD094667 (NICHD)). Dr. Ackerman is supported by NIH/K08DK118176, NIH/R03AG067993, DOD W81XWH2110644.

**Conflicts of Interest:** Dr. Ackerman is a consultant for Watershed Medicaland Abbvie, and an investigator for MicrogenDx and Medtronic.

### Competing Interest Statement

Dr. Ackerman is a consultant for Watershed Medicaland Abbvie, and an investigator for MicrogenDx and Medtronic

### Funding Statement

Funding: This work was supported by the AUGS/Duke UrogynCREST (Urogynecology Clinical Research Educational Scientist Training) Program (R25HD094667 (NICHD)). A.L.A. is supported by NIH/K08DK118176, NIH/R03AG067993, DOD W81XWH2110644.

### Author Declarations

This study was approved by the Cedars-Sinai Medical Center Institutional Review Board (IRB#00040261).

