## Supplementary figures and images for "Myofascial Frequency Syndrome: A novel syndrome of bothersome lower urinary tract symptoms associated with myofascial pelvic floor dysfunction"

### Supplemental Figure 1

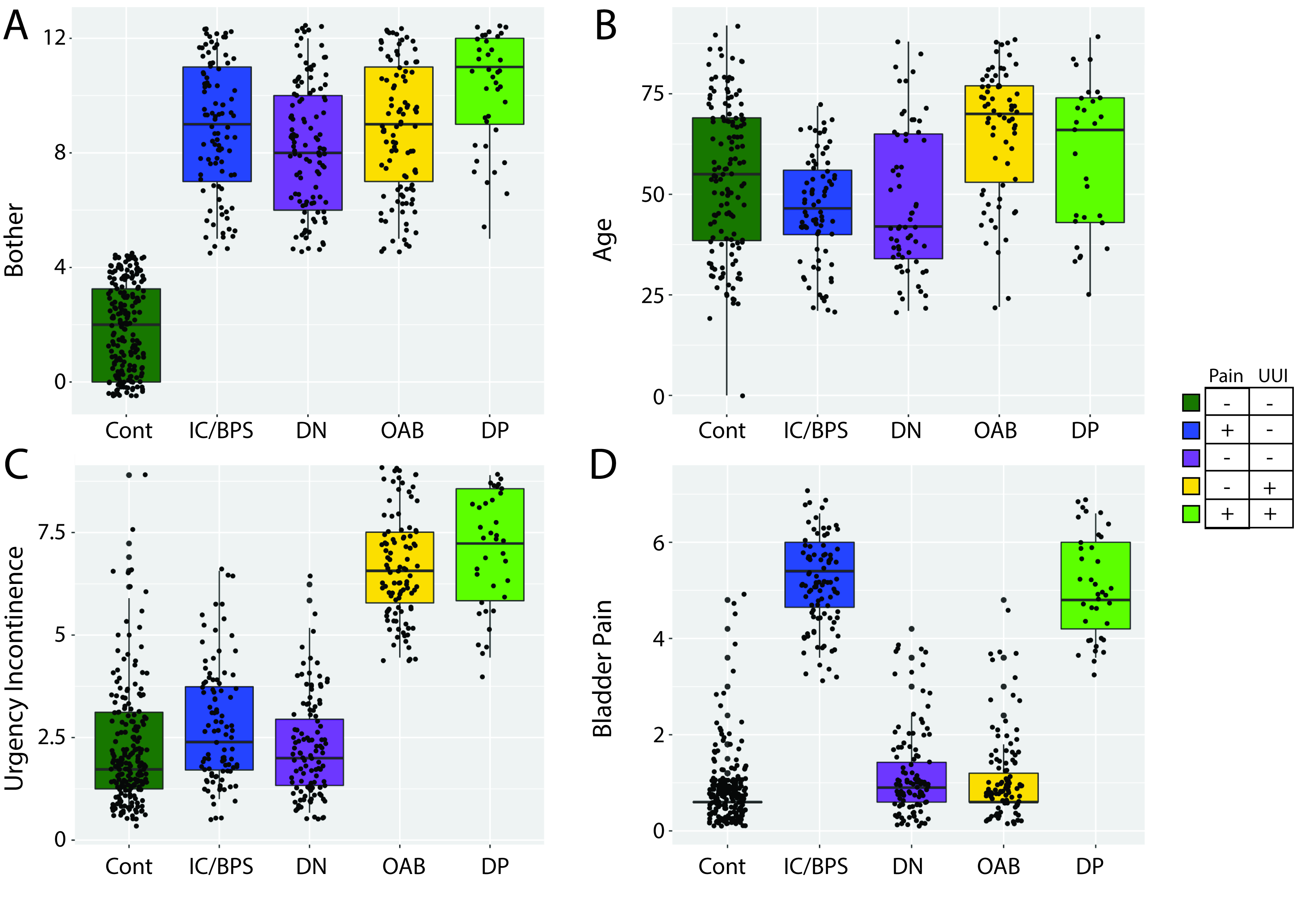
